# Zika Virus looming epidemic in Pakistan: seroprevalence findings by plaque reduction neutralization test in the Sindh Region of Pakistan

**DOI:** 10.1101/2023.04.27.23289241

**Authors:** Khekashan Imtiaz, Joveria Farooqi, Kelli L. Barr, Akbar Kanji, Dhani Prakoso, Zahida Azizullah, Maureen T. Long, Erum Khan

## Abstract

**Background:** Zika virus (ZIKV) has emerged as a cause of febrile illness in children and adults globally. West Asian and Middle Eastern countries have not yet experienced the widespread emergence of ZIKV. In Pakistan, detection of ZIKV antibodies have been reported. However, the validity of this data is questionable given the current understanding of flaviviral antigenic cross-reactivity. In order to determine if ZIKV is circulating in the Sindh region of Pakistan, patients presenting to healthcare centers with an acute febrile dengue-like illness were evaluated for ZIKV infection.

**Methods:** Dengue virus (DENV) screening in patients was performed using a commercial ELISA Rapid Test NS1 antigen capture test. All DENV negative samples were tested for Zika virus, using a commercial IgM capture ELISA kit. Additionally, a plaque reduction neutralization test (PRNT) was performed to measure neutralizing antibodies. Singleplex, two-step Real-time PCR using general primers and probes was performed for the detection of Zika virus nucleic acid.

**Results:** Patients with acute dengue-like illness (DLI) presenting at healthcare centers in different sites throughout the Sindh region of Pakistan were recruited. A total of 745 patient samples were tested for ZIKV via IgM ELISA and RT-PCR. Thirty-seven samples (4.9%) tested positive for ZIKV IgM without any cross-reactivity to other flaviviruses tested simultaneously. These were considered as presumptive positive for ZIKV, selected presumptive positive samples (n=20) were confirmed using PRNT50 using Vero cells. All 20 samples showed ZIKV neutralization at PRNT50.

**Conclusions:** Our study provides evidence that ZIKV is circulating in the Sindh region of Pakistan and is a probable cause of clinical dengue-like illness (DLI) cases that are seen seasonally in Pakistan.

## Introduction

Zika virus (ZIKV) has emerged as a cause of febrile illness in children and adults globally. West Asian and Middle Eastern countries have not yet experienced the widespread emergence of ZIKV. In Pakistan, detection of ZIKV antibodies has been reported, however, the validity of this data is questionable given the current understanding of flaviviral antigenic cross-reactivity(1).

The Zika virus (ZIKV) is ∼40–60 nm enveloped and has an icosahedral shell embedded with surface projections. The genome of ZIKV contains a non-segmented, positive-sense, single-stranded RNA of about 11 kb that encodes around 3500 amino acids and has a single long open reading frame (ORF) (5′-C-prM-E-NS1-NS2A-NS2B-NS3-NS4A-NS4B-NS5-3′) with flanking untranslated (UTR) regions at both ends(2). The genome has been shown to encode a single polypeptide, which is cleaved by host-cell proteases into three structural proteins (capsid (C), envelope (E), and precursor membrane (prM) or membrane (M)) and seven nonstructural proteins (NS1, NS2A, NS2B, NS3, NS4A, NS4B, and NS5)(3).

In the past years, outbreaks of DENV and dengue-like illness (DLI) has affected large proportions of the Pakistani population. Chikungunya virus (CHIKV) has recently been confirmed in Karachi which is the largest metropolitan city in Pakistan(4). Arbovirus infections are often misdiagnosed and overlooked because of unclear clinical symptoms and extensive differential diagnose that overlap with other viral pathogens. Although, no confirmed case of ZIKV has been identified in Pakistan, however, serological evidence exists(5). One of the possible reasons that might obscure the detection of ZIKV is the lack of a proper screening system that can differentiate ZIKV from other flaviviruses. The laboratory confirmation of ZIKV is challenging because of serological cross-reactivity with other flaviviruses, particularly with DENV(6). There is a high probability that ZIKV is being transmitted in Pakistan but is unreported or misdiagnosed because many people do not visit doctor due to mild symptoms. Further, ZIKV shares not only its vector *Aedes aegypti* with CHIKV and DENV but also the clinical features of disease and geographic distribution(7).

Published studies, have shown that ZIKV co-circulates with other flaviviruses, including DENV, West-Nile virus (WNV), Japanese Encephalitis virus (JEV), and Yellow Fever virus (YFV)(8). Most of the evidence is based on sero-surveillance. Antibody-based assays can be problematic as serological cross-reactivity can complicate results(9). Furthermore, previous exposures and co-infections can further complicate diagnostic tests(10). Additional difficulties arise when diagnosing clinical samples in different locations as reference virus and antigen sources can produce divergent results in response to locally circulating viral isolates(11). Unfortunately, no such studies were reported from Pakistan.

The aim of this study is to investigate the presence of ZIKV as cause of acute febrile illness in patients presenting with DLI in different regions of Sindh province of Pakistan by IgM ELISA, RT-PCR and confirmation by PRNT.

## Material and Methods

### Ethics statement

The study protocol was approved by Ethics Review Committee, Aga Khan University, Karachi, Pakistan (#3183-PAT-ERC-14). All enrolled subjects gave written informed consent in accordance with the Declaration of Helsinki. Patients with history of fever for 5-7 days were approached through previously trained health professional to explain the importance of study. Patients were recruited only after their formal consent was taken.

### Study sites

Five study sites were established in the Sindh province in Pakistan, and research personnel were trained for demographic and clinical data and sample collection. These sites included five hospitals: Aga Khan University Hospital (Karachi), Ghulam Mohammad Mahar Medical College (Sukkur), CMC Teaching Hospital (Larkana), Muhammed Medical College Hospital (Mirpurkhas), and the Civil Hospital of Hyderabad, Pakistan. (Figure 2)

**Figure 1.**
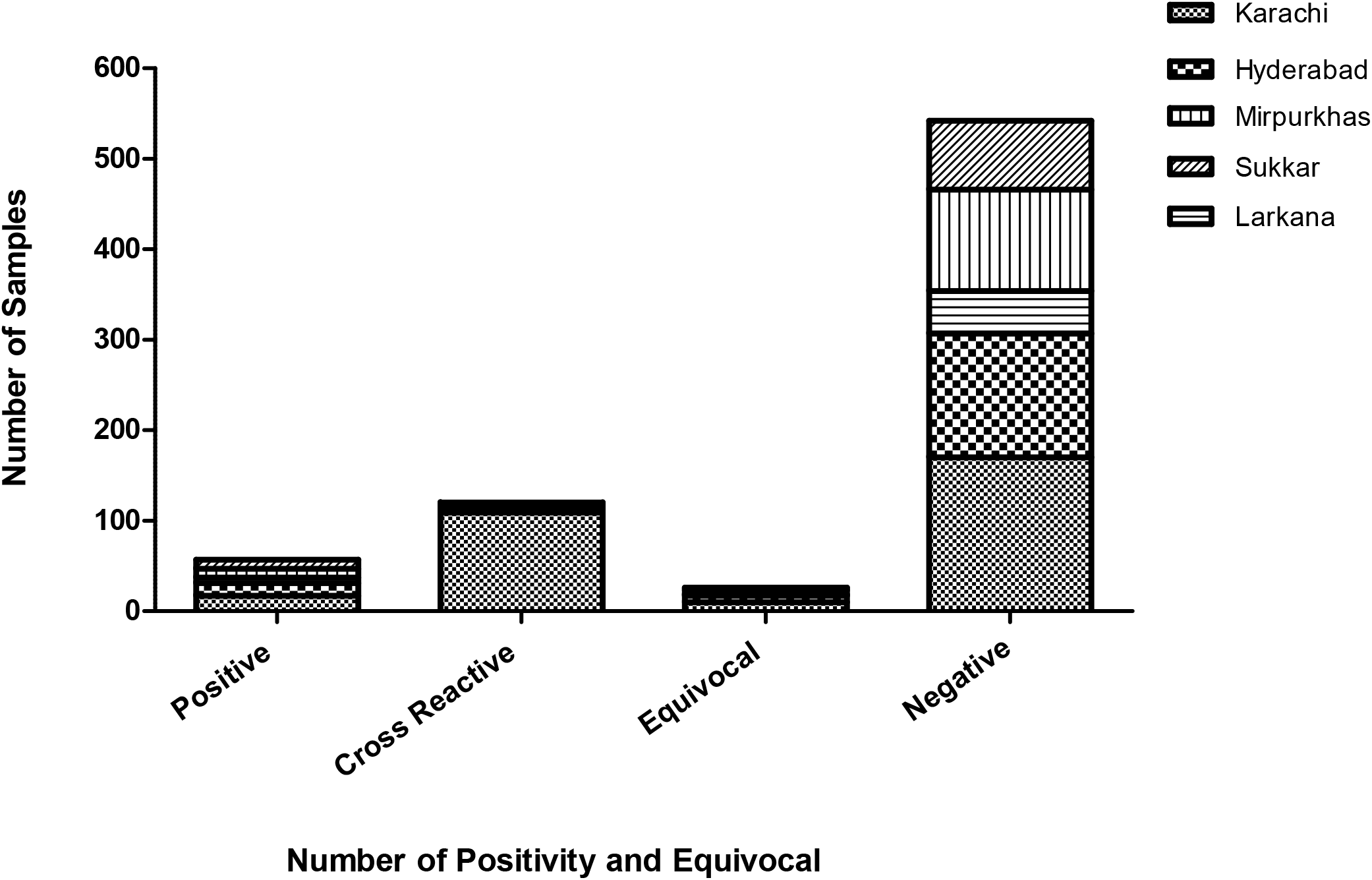
Depicts the Zika virus IgM reactivity of patients (n=745) presenting with acute febrile illness from five regions of Sindh Province of Pakistan 2015-17

### Inclusion and Exclusion Criteria

All patients including males and females between 10 and 65 years of age meeting the case definition on the day of enrollment, were eligible for participation in the study. Patients younger than 10 and older than 65 years of age and those who tested positive for CCHF, influenza, malaria, tuberculosis, and bacterial septicemia during routine hospital admittance procedures were excluded.

### Demographic and Clinical Characteristics of Patients

A cross-sectional, observational study was conducted from April 2015 to June 2017. Using convenience-based sampling seven hundred and forty-five serum samples were collected to determine ZIKV infection and/or expose. The following demographic and clinical variables were evaluated: sex, age, enrolment site and presence of fever, rash, myalgia, arthralgia, headache, conjunctivitis/ eye pain, nausea, diarrhea, and neurological symptoms like paranesthesia, muscle weakness. The laboratory parameters investigated were: complete blood count (CBC), aspartate aminotransferase (AST), alanine aminotransferase (ALT). Lymphopenia was defined as the total lymphocyte count under 1000 per mm^3^.

The presence of ZKIV IgM and neutralizing antibodies was considered as presumptive positive case for zika virus as defined by CDC guidelines(12).

The 20.4% (n=152) showed cross reactivity with Dengue, West Nile, Japanese Encephalitis and Chikungunya viruses, while 74.6% (n=556) samples were negative of DENV NS1 antigen and IgM ELISA

### Serology

Patient samples were screened using a commercial DENV (Panbio Dengue Early Rapid Test NS1 antigen capture test, Alere, Waltham, MA) following the manufacturer’s instructions. Samples reported negative for Dengue NS1 Ag, were tested for ZIKV using commercial IgM capture ELISA (ZIKV Detect™, InBios, Seattle, WA) following the manufacturer’s instructions. According to manufacturer’s guideline a Zika immune status ratio (ZIKA ISR value ≥ 1.70) considered presumptive positive.

### Real-Time PCR (RT PCR)

Total RNA was extracted from 140μl of the patient serum samples using the QIAamp Viral RNA Mini Kit (Qiagen, Germany) according to the manufacturer’s protocol(10). The RNA was eluted in 60ul of nuclease-free water and stored at -80^0^C until needed for detection of nucleic acids ZIKV primer sequences were constructed for possible strains circulating in Pakistan via addition of degenerate nucleotides. RT-PCR was performed using a commercial master mix (BioRad iTaq Universal Probes Supermix, BioRad, Hercules, CA, USA) on a BioRad CFX96 Real Time PCR machine(13).

RT-PCR for ZIKV isolates was performed using the two-step RT-PCR protocol with ZIKV general primers and probe (22, 26). The probe was modified with an internal Nova quencher, 5’ FAM-AGCCTACCT[Nova]TGACAAGCAGTCAGACACTCAA BHQ-1 3’ to improve sensitivity(13). WNV and JEV primers and probes were used to detect the NS2 gene(14). DENV primers and probes were used to detect the NS5, envelope, or prM protein depending on serotype(13).

#### Cell Cultures

Vero cells (ATCC CCL-81) were grown in Dulbecco Minimal Eagles Medium (DMEM) (Sigma) supplemented with 10% Fetal Bovine Serum (FBS), 1% L-glutamine 200mM, 1% penicillin G (100U/ml), streptomycin (100ug/ml) at 37°C and 5% CO_2._

#### Virus Stock

ZIKV strain was obtained from University of Texas Medical Branch (UTMB). The viral stock was prepared by inoculation of Vero cells contained in 75cm^2^ culture flasks with virus diluted in 1ml of DMEM containing 5% FBS. After 1hour, 14ml of DMEM supplemented with 10% FBS was added, and the cells were cultured for 5 to 7 days. Cell culture supernatant was then harvested and centrifuged at 4000rpm for 5 min to removed cell debris. To observe, ZIKV load, RT-PCR was performed at Days 0, 3 and 5 to verify viral load. The supernatant containing the virus was adjusted to 20% FBS, aliquoted, and stored at −80^0^C.

### Plaque Reduction Neutralization Test (PRNT)

For this study (PRNT) was adapted from previously described protocols(14). It was carried out with ZIKV control strain obtained from (University of Texas Medical Branch, UTMB) diluted to 50pfu/20ul to give 2500pfu/ml in 12 well plates containing confluent Vero cell monolayer. After 1-hour incubation at 37^0^C in 5% CO_2_ incubator, each 4-fold serum dilution (1:10, 1:40 and 1:160) and virus mixture was transferred onto confluent Vero cells monolayer in 12 well plates. Following incubation for 1hr at 37^0^C in 5% CO_2_ incubator, serum and virus inoculum was removed and cell monolayers was immediately covered with 1.0% carboxymethylcellulose (CMC)/DMEM overlay. Plaques were counted at day 5 post-infection (pi). Serum specimens with 50% (PRNT50) reduction of the number of plaques at titers ≥10 were recorded as positive according to the recommendation of Centers for Disease Control and Prevention(15).

### Statistical analysis

Statistical analysis was performed using Stata software version 12.01 and the chi-square test or Fisher’s exact test was used to verify the association between categorical variables was performed to compare percentages of IgM ELISA positive, cross reactive and negative Zika virus cases and p<0.05 was found to be significant.

## Results

### Serological evidence of ZIKV amongst febrile patients in Sindh

A total of 745 patients with undifferentiated acute febrile illness presenting to tertiary care center were assessed for ZIKV between April 2015 -17. The 4.9 % patients (n=37) tested positive for IgM antibodies, without any cross reactivity to other flaviviruses tested simultaneously, which included DENV, JEV, and WNV. These patients were considered presumptive IgM positive for ZIKV infection as per CDC guidelines(16). Among the cross-reactive samples, 20.4% (n=152) showed cross reactivity to other tested flaviviruses, notably DENV (13.9%), WNV (14%), and JEV (11.9%).

The 20 serum samples of patients identified as presumptive positive on the basis of high IgM titers on ELISA, were selected for neutralization studies using Vero cell line. A plaque reduction neutralization assay (PRNT_50_) a titer reciprocal of the endpoint serum dilution that reduces the challenge of DENV, JEV, and WNV viruses, plaque count by 50% was performed. All 20 samples showed neutralization (PRNT50) of Vero cells infected with the ZIKV.

### Patient Characteristics

Based on MAC-ELISA IgM test results, patients were categorized as ZIKV seropositive (n=37), cross-reactive (n=153) and seronegative (n=556). The demographic and clinical presentations of ZIKV seropositive, cross-reactive, and seronegative cases are shown in Table 1. The probability of cross-reactivity in children was less as compared to adults. No propensity in age group for seropositivity was observed. There was no gender predilection for ZIKV seropositivity. However, there was a greater likelihood of cross-reactivity in males. Out of 745 patients recruited, four hundred and nine (54.89%) were females and three hundred and thirty-six (45.10%) were males. Compared to urban regions like Karachi and Hyderabad, the seropositivity for ZIKV was higher in patients from rural regions such as Larkana, Mirpurkhas, and Sukkar. Within Sindh, the cross-reactivity was highest in Karachi compared to the other cities of the province.

**Table 1:** Demographic and Clinical presentation of patients presenting with acute febrile illness compared with ZIKV IgM reactivity from Sindh Pakistan 2015-17.

| Characteristic | Total N | Positive n(%) | Cross Reactive n (%) | Negative n (%) | Chi-sq pvalue |
| --- | --- | --- | --- | --- | --- |
| **Age group** |  |  |  |  | 0.019 |
| <=18 | 176 | 11 (6.3) | 22 (12.5) | 143 (81.3) |  |
| >18-30 | 268 | 14 (5.2) | 72 (26.9) | 182 (67.9) |  |
| 31-60 | 260 | 11 (4.2) | 64 (24.6) | 185 (71.1) |  |
| >60 | 41 | 1 (2.4) | 8 (19.5) | 32 (78.0) |  |
| **Sex** |  |  |  |  | 0.012 |
| Male | 336 | 12(3.5) | 90(26.7) | 234(69.6) |  |
| Female | 409 | 25(6.1) | 76(18.5) | 308(75.3) |  |
| **Field Sites** |  |  |  |  |  |
| Karachi | 306 | 10(3.2) | 126(41.1) | 170(55.5) | <0.001 |
| Hyderabad | 163 | 6(3.6) | 20(12.2) | 137(84.0) |  |
| Larkana | 55 | 5(9.0) | 3(5.4) | 47(85.4) |  |
| Mirpurkhas | 131 | 9(6.8) | 10(7.6) | 112(85.5) |  |
| Sukkar | 90 | 7(7.7) | 7(7.7) | 76(84.4) |  |
| **Clinical Presentations** |  |  |  |  |  |
| Fever | 689 | 36 (5.2) | 160 (23.2) | 493 (71.5) | 0.035 |
|  | 56 | 1 (1.8) | 6 (10.7) | 49 (87.5) |  |
| Headache | 499 | 29 (5.81) | 75 (15.0) | 395 (79.1) | <0.001 |
|  | 246 | 8(3.2) | 91(36.9) | 147(59.7) |  |
| Bodyache | 431 | 20(4.6) | 81(18.7) | 330(76.5) | 0.05 |
|  | 314 | 17(5.4) | 85(27.0) | 212(67.5) |  |
| Arthralgia | 200 | 10(5.0) | 26(13.0) | 164(82.0) | 0.001 |
|  | 545 | 27(4.9) | 140(25.6) | 378(69.3) |  |
| Nausea | 147 | 5(3.4) | 56(37.1) | 86(58.5) | <0.001 |
|  | 598 | 32(5.3) | 110(18.3) | 456(76.2) |  |
| Rash (allergy/ maculopapular rash) | 5 | 1(20.0) | 3(60.0) | 1(20.0) | 0.013* |
|  | 740 | 36(4.8) | 163(22.0) | 541(73.1) |  |
| Eye Pain | 175 | 9(5.1) | 17(9.7) | 149(85.1) | <0.001 |
|  | 570 | 28(4.9) | 149(26.1) | 393(68.9) |  |
| CNS (Neurological symptoms) | 313 | 16(5.1) | 25(7.9) | 272(86.9) | <0.001 |
|  | 432 | 21(4.8) | 141(32.6) | 270(62.5) |  |

The clinical presentation of patients seropositive for ZIKV, as represented in Table 1, was more likely if the patient was febrile. However, the overlap with other non-specific symptoms like headache, body ache, vomiting, and nausea was prominent in all three groups. There was no significant association of ZIKV seropositivity with arthralgia, eye pain or neurological symptoms. The number of patients with rash were too few to come to any plausible conclusion. However, the Fisher Exact test did show significance for seropositivity and cross-reactivity with ZIKV (p value 0.013).

Laboratory findings of patients are presented in Table 2. The review of the Complete blood count (CBC), showed that the proportion of leucopenia and monocytopenia was low in ZIKV seropositive cases, while there was a comparatively higher occurrence of these findings in cross-reactive cases. Furthermore, there was a significant association between cross-reactivity and thrombocytopenia, and significantly less cross-reactivity with lymphopenia and neutrophilia. There was no significant association of ZIKV seropositivity with eosinophilia or eosinopenia, but significantly less proportion of eosinophilia findings with cross-reactive cases. The rise in ALT and AST was not associated with ZIKV seropositivity, however, there was a significantly higher propensity for cross-reactivity when these enzyme levels were raised.

**Table 2:** Laboratory findings of patients presenting with acute febrile illness compared with ZIKV IgM reactivity from Sindh Pakistan 2015-17.

| Characteristics | Total N=745 | Positive<br>n=37(4.9%) | Cross Reactive n=152<br>(20.4%) | Negative n=556<br>(74.6%) | P Value |
| --- | --- | --- | --- | --- | --- |
| AST | 104 | 2(1.9) | 65(62.5) | 37(35.5) | <0.001 |
|  | 75 | 4(5.3) | 14(18.6) | 57(76.0) |  |
| ALT | 113 | 2(1.7) | 64(56.6) | 47(41.5) | <0.001 |
|  | 135 | 9(6.6) | 39(28.8) | 87(64.4) |  |
| Anemic | 426 | 23(5.4) | 64(15.0) | 339(79.5) | <0.001 |
|  | 319 | 14(4.3) | 102(31.9) | 203(63.6) |  |
| Leucopenia | 143 | 2(1.4) | 91(63.6) | 50(34.9) | <0.001 |
| Leukocytosis | 136 | 7(5.1) | 8(5.8) | 121(88.9) |  |
|  | 466 | 28(6.0) | 67(14.3) | 371(79.6) |  |
| Thrombocytopenia | 228 | 8(3.5) | 116(50.8) | 104(45.6) | <0.001 |
| Thrombocytosis | 75 | 6(8.0) | 5(6.6) | 64(85.3) |  |
|  | 442 | 23(5.2) | 45(10.1) | 374(84.6) |  |
| Lymphopenia | 232 | 13(5.6) | 28(12.0) | 191(82.3) | <0.001 |
| Lymphocytosis | 82 | 3(3.6) | 38(46.3) | 41(50.0) |  |
|  | 431 | 21(4.8) | 100(23.2) | 310(71.9) |  |
| Neutrophilia | 164 | 9(5.4) | 17(10.3) | 138(84.1) | <0.001 |
| Neutropenia | 76 | 2(2.6) | 35(46.0) | 39(51.3) |  |
|  | 505 | 26(5.1) | 114(46.0) | 365(72.2) |  |
| Eosinophilia | 28 | 2(7.1) | 2(7.1) | 24(85.7) | <0.001* |
| Eosinopenia | 340 | 15(4.4) | 100(29.4) | 225(66.1) |  |
|  | 377 | 20(5.3) | 64(16.9) | 293(77.7) |  |
| Monocytosis | 149 | 4(2.6) | 47(31.5) | 98(65.7) | 0.024* |
| Monocytopenia | 13 | 1(7.6) | 3(23.0) | 9(69.2) |  |
|  | 583 | 32(5.4) | 116(19.9) | 435(74.6) |  |
| Basophilia | 1 | 0(0.0) | 1(100.0) | 0(0.0) | 0.272* |
|  | 735 | 36(4.9) | 163(22.1) | 536(72.9) |  |
| MP test | 0 | 0(0.0) | 0(0.0) | 6(100.0) | 0.518* |
|  | 739 | 37(5.0) | 166(22.4) | 536(72.5) |  |
| BLCS | 33 | 3(9.0) | 3(9.0) | 27(81.8) | 0.21 |
|  | 609 | 31(5.0) | 123(20.2) | 455(74.7) |  |

## Discussion

Confirmation of ZIKV diagnosis in a region endemic with other flaviviruses like DENV is challenging due to flavivirus cross reactive antibodies(17). Therefore, MAC-ELISA results in dengue endemic region must be confirmed by a highly specific test like PRNT. In this study, we report the confirmation of ZIKV IgM antibodies with viral neutralization antibodies in serum samples of patients that were seropositive for ZIKV at titer PRNT50. Clinical specimens were confirmed positive based on the CDC guidelines that require detection of virus-specific IgM antibodies with virus-specific neutralizing antibodies in the same or later specimen(17). We found that InBios the MAC-ELISA IgM assay using the ZIKV envelope glycoproteins, had good sensitivity and specificity allowing assessment of differential detection of ZIKV antibodies. We were able to detect patients that were ZIKV seropositive only (n=37), cross-reactive (n=153) and seronegative (n=556). Of the ZIKV seropositive only group, we were able to show viral neutralization using via PRNT at titer PRNT50. To the best of our knowledge this is the first confirmed report of Zika virus from Pakistan.

ZIKV has been circulating since at least the 1960s in several countries of the South-East Asia Region (SEAR)(18). In spite of this long history of ZIKV circulation in the region, information on the epidemiology of ZIKV infection and its associated complications in the SEAR remain limited. Prior to 2015, cases of ZIKV infection in Pakistan were identified in residents and returning travelers from Bangladesh, Indonesia, the Maldives, and Thailand confirmed by serological and/or molecular testing (19-25). Detection of congenital syndrome (CGS) has been limited in the SEAR, mostly due to low levels of transmission in the general population or limited epidemiological data. Improved surveillance and epidemiologic investigations are needed to better ascertain the incidence of ZIKV infection in the SEAR and its impact on birth outcomes. According to the World Health Organization (WHO) 2019 report, a total of 87 countries and territories have had evidence of mosquito-borne transmission of (ZIKV), distributed across four of the six WHO Regions (African Region, Region of the Americas, South-East Asia Region, and Western Pacific Region). Despite, the limited epidemiologic data from the WHO regions, including Pakistan, new scientific evidence continues to increase our understanding of global ZIKV transmission and its associated complications(1).

Analysis of climate, previous flaviviruses outbreaks, traveling patterns, and mosquito biology has improved our ability to predict the expected path and next landing spots of ZIKV in Pakistan. For example, an epidemic of ZIKV is expected to occur between July and September as monsoon rainfall provides suitable breeding habitats for mosquitoes. People living in slum areas of Pakistan are at greater risk of infection due to standing water and open sewage drains that are breeding sites for the mosquito(26).

It is anticipated that Karachi and Lahore, the two largest cities of Pakistan are more susceptible to ZIKV outbreaks because of rapid urbanization, growth of slum areas and vector breeding grounds. Besides Karachi and Lahore, other metropolitan areas like Islamabad, Multan, Quetta, Gwadar, and Peshawar are potential routes of ZIKV entry because of major airline traffic between epidemic regions(26).

Studies have shown that ZIKV can be detected by RT-PCR in urine for 10–14 days after symptom onset but only for approximately 3 days from serum(27). It has been reported that when undetected by RT-PCR, diagnosis is made following guidelines that require the presence of both virus-specific IgM antibodies and virus-specific neutralizing antibodies(12). Whilst clinicians use IgM ELISA to screen for infection, the plaque reduction neutralization test (PRNT) with ZIKV alongside DENV and other endemic flaviviruses is recommended by the Centers for Disease Control and Prevention (CDC) to confirm diagnosis in RT-PCR negative and IgM ELISA positive cases(15). However, PRNT has been shown to be challenging for diagnosis with over half of patients exhibiting significant neutralization of ZIKV, DENV, and other flaviviruses co-circulating in that region(28).

This study employed a commercial assay for the detection of ZIKV-specific IgM antibodies. The internal flaviviral control in many commercial ZIKV IgM kits is an important addition for normalizing out cross-reactive background. We found the InBios assay effective in differentiating cross reactive and non-reactive samples from true positive specimen, that were further confirmed for ZIKV by PRNT.

The PRNT plays an important role in confirming ZIKV infections, as well as in identifying false-positive ZIKV IgM results to rule out ZIKV infection. Previous studies have shown serologic cross-reactivity between ZIKV and related flaviviruses, including DENV, but the degree to which neutralizing antibody cross-reactivity limits the ability to identify the specific virus responsible for the current infection is not clear(29). One published report suggests that relative levels of neutralizing antibody titers can distinguish ZIKV from DENV infections, especially in specimens collected months after infection(30). In our study we did find patients that were positive only for ZIKV that did not show any cross reacting IgM for DENV and these also showed neutralizing effects against ZIKV at PRNT50.

Detection of ZIKV RNA in serum is considered the confirmatory test for diagnosis of ZIKV infection in both symptomatic and non-symptomatic cases. One of the challenges is the timing at which the serum sample is collected, the viremia is high during the first few days of acute febrile phase of infection(6). In this study, we were not able to detect ZIKV RNA in the serum samples of the patients although most of the serum samples were collected during days 5-7 of febrile illness. It has been shown that ZIKV nucleic acid detection is lowest in serum however the testing sensitivity can be maximized up to 2 weeks post-infection by testing whole blood and urine samples simultaneously(31).

A limitation of this study is that PRNT50 was performed on small number of patients (n=20). The study was only performed on patients negative for DENV, hence patients co-infected with DENV and ZIKV could not be identified. Additionally, testing could not be extended to PRNT90 due to limited funds.

Additionally, we did not collect urine samples, as its testing by PCR requires special considerations such as time of collection, temperature of sample during transport and storage. Storage of urine at 4°C for periods longer than 48 hours is considered to affect ZIKV RNA detection by RT-PCR, additionally freezing at −80°C may result in significant loss of detectable ZIKV RNA in low positive samples(32). Because of these limitations, the RT-PCR testing was only performed on serum samples which is one of the possible limitations of this study.

The laboratory findings show that the majority of patients presented with the normal CBC findings (78.26%), and up to 76% did not show elevated levels of AST and ALT liver enzymes. These findings may help differentiate ZIKV-infected patients from those with DENV where the liver enzymes are often elevated and blood CBC is often significant for elevated hematocrit, monocytosis and decreased platelet, and may assist in developing a clinical diagnosis(33).

## Conclusions

Our study provides evidence that ZIKV is circulating in Sindh region of Pakistan as population is showing seroconversion to this virus. It is a probable cause of clinical dengue-like illness (DLI) cases that are seen seasonally in Pakistan. This is the first study to confirm patients with serum IgM antibodies for ZIKV using PRNT assay. Timely testing of serum and urine samples simultaneously by molecular techniques must be employed in patients presenting with DLI to further confirm these findings in Pakistan

## Data Availability

All the data is present within the manuscript

## Author Contributions

The following authors contributed to this manuscript in the following ways: contribution to the conception and design of the work: KB, EK, KI, JF, AK, DP and ML and acquisition, analysis, and interpretation of data: EK, KB, JF, AK, KI and ML. Drafting, editing, revising, and approving drafts: KB, EK, KI, JF, AK, DP and ML. All agreed to be accountable for all aspects of the work.

## Acknowledgments

We are thankful to World Reference Center for Emerging Viruses and Arboviruses (WRCEVA) for sharing isolates of ZIKV; Sally Beachboard, for negotiating costs of supplies for our work in both the USA and Pakistan, University of Florida, Gainesville.

